# Same-day testing with initiation of antiretroviral therapy or tuberculosis treatment versus standard care for persons presenting with tuberculosis symptoms at HIV diagnosis: A randomized unblinded trial

**DOI:** 10.1101/2022.12.28.22283999

**Authors:** Nancy Dorvil, Vanessa R. Rivera, Cynthia Riviere, Richard Berman, Patrice Severe, Heejung Bang, Kerlyne Lavoile, Jessy G. Devieux, Mikerlyne Faustin, Giovanni Saintyl, Maria Duran Mendicuti, Samuel Pierre, Alexandra Apollon, Emelyne Dumond, Guyrlaine Pierre-Louise Forestal, Vanessa Rouzier, Adias Marcelin, Margaret L. McNairy, Kathleen F. Walsh, Kathryn Dupnik, Lindsey K. Reif, Anthony L. Byrne, Stephanie Bousleiman, Eli Orvis, Patrice Joseph, Pierre-Yves Cremieux, Jean William Pape, Serena P. Koenig

## Abstract

**Background:** Same-day HIV testing and antiretroviral therapy (ART) initiation is being widely implemented. However, the optimal timing of ART among patients with tuberculosis (TB) symptoms is unknown. We hypothesized that same-day treatment would be superior to standard care in this population.

**Methods and Findings:** We conducted an open-label randomized trial among adults with TB symptoms at initial HIV diagnosis at GHESKIO in Haiti. Participants were randomized in a 1:1 ratio to same-day treatment (same-day TB testing with same-day treatment [TB medication if TB; ART if no TB]) vs. standard care. In both groups, ART was initiated two weeks after TB treatment. The primary outcome was retention in care with 48-week HIV-1 RNA <200 copies/mL, with intention to treat analysis. From November 6, 2017 to January 16, 2020, 500 participants were randomized (250/group). Baseline TB was diagnosed in 40 (16.0%) in the standard and 48 (19.2%) in the same-day group; all initiated TB treatment. In the standard group, 245 (98.0%) initiated ART at median of 9 days; 6 (2.4%) died, 229 (91.6%) were retained, and 220 (88.0%) received 48-week HIV-1 RNA testing; 168 had <200 copies/mL (among randomized: 67.2%; among tested: 76.4%). In the same-day group, 249 (99.6%) initiated ART at median of 0 days; 9 (3.6%) died, 218 (87.2%) were retained, and 211 (84.4%) received 48-week HIV-1 RNA; 152 had <200 copies/mL (among randomized: 60.8%; among tested: 72.0%). There was no difference between groups in the primary outcome (60.8% vs. 67.2%; risk difference: -0.06; 95% CI: -0.15, 0.02; p=0.14). The main limitation of this study is that it was conducted at a single urban clinic, and the generalizability to other settings is uncertain.

**Conclusions:** In patients with TB symptoms at HIV diagnosis, same-day treatment is not associated with superior retention and viral suppression. A short delay in ART initiation, which facilitates more feasible TB testing, does not compromise outcomes.

**Trial Registration:** This study is registered with ClinicalTrials.gov NCT03154320

## INTRODUCTION

Initiating antiretroviral therapy (ART) on the day of HIV diagnosis (same-day ART) or as soon as possible afterwards is now recommended in treatment guidelines worldwide, with the goal of improving ART uptake and linkage to care and decreasing time to viral suppression (1-3). However, a substantial proportion of patients worldwide report tuberculosis (TB) symptoms at HIV diagnosis, and the optimal timing of ART initiation in this group of patients is unknown (4).

The 2017 World Health Organization (WHO) guidelines stated as a clinical consideration that ART should be briefly delayed while investigating for TB (5). In 2021 the WHO guidelines were amended to recommend same-day ART initiation, while TB test results are pending, with close follow-up to initiate TB treatment if a diagnosis of TB is made. This new WHO recommendation was based on the concern that delays in ART initiation in patients with TB symptoms may increase rates of pretreatment lost to follow-up (LTFU) (1). However, the WHO guidelines also state that very little information is available on the potential harm of same-day ART initiation in the presence of TB symptoms (6). Potential adverse events of ART initiation in the presence of untreated TB include pulmonary dysfunction and immune reconstitution inflammatory syndrome (IRIS) (7, 8). A recent systematic review found there was insufficient evidence about whether the presence of TB symptoms at HIV diagnosis should lead to ART deferral (9).

It is logistically challenging to complete a TB evaluation on the same day of HIV diagnosis, especially in lower-income settings, as the Xpert MTB/RIF assay (Cepheid, Sunnyvale, CA) has at least a two-hour turnaround time, and tests are often conducted in centralized laboratories, with further delays for specimen transport, testing backlog, and communication of test results. Furthermore, the sensitivity of one Xpert MTB/RIF testing is about 80% in persons living with HIV (PLWH), and a single negative test does not rule out active TB (10).

A short delay to conduct TB testing is more feasible than same-day TB testing prior to ART initiation in many settings. However, data on the consequences of short delays in ART initiation are limited. In the clinical trials of same-day ART vs. standard care, the study populations either excluded or included a minority of patients with TB symptoms (11-14). Furthermore, these trials were conducted several years ago, when the standard of care included multiple visits prior to ART initiation. Consequently, participants in the comparison groups generally initiated ART weeks to months after HIV diagnosis, while the current standard of care has shifted to avoid delays in ART initiation.

We conducted a randomized trial to determine whether same-day TB testing and treatment initiation (TB medication or ART) was associated with superior outcomes, compared with standard care (initiating TB treatment within 7 days and delaying ART to Day 7 if TB not diagnosed), in adults with TB symptoms at HIV diagnosis. We hypothesized that same-day treatment would improve the primary outcome of retention with viral suppression at 48 weeks after enrollment.

## METHODS

### Study Design and Setting

We conducted an unblinded, randomized trial of same-day treatment vs. standard care for patients who presented with symptoms of TB at HIV diagnosis at the Haitian Group for the Study of Kaposi’s Sarcoma and Opportunistic Infections (GHESKIO) in Port-au-Prince, Haiti. The primary study endpoint was the proportion of participants in each group with 48-week HIV-1 RNA <200 copies/mL.

GHESKIO is a Haitian non-governmental organization and the largest provider of HIV and TB treatment in the Caribbean. HIV care at GHESKIO is largely funded by the US President’s Emergency Plan for AIDS Relief (PEPFAR) and the Global Fund to Fight AIDS, Tuberculosis, and Malaria. All care is provided free of charge. The HIV prevalence in Haiti is an estimated 1.8% and the incidence of TB is 168/100,000 (15, 16). The study was approved by the institutional review boards at GHESKIO, Mass General Brigham, Florida International University, and Weill Cornell Medical College.

### Study Participants

Participants were recruited if they tested positive for HIV at GHESKIO and reported at least one TB symptom on a routinely conducted questionnaire. Patients were eligible for study inclusion if they were infected with HIV-1, ≥18 years of age, able to provide informed consent, and reported cough, fever, and/or night sweats of any duration and/or weight loss, which was confirmed by the study physician. Initially enrollment was limited to patients who reported loss of at least 10% of body weight, but in April 2018, this was changed to any reported weight loss, because most patients were unaware of their exact change in weight.

Patients were excluded if they had received ART previously, failed to demonstrate preparedness on an ART readiness questionnaire, were treated for TB in the prior year, were pregnant or breastfeeding, had active drug or alcohol use, or a mental condition that would interfere with their ability to adhere to study requirements, or planned to transfer care during the study period. Patients were also excluded if they presented with symptoms consistent with WHO stage 4 neurologic disease (cryptococcal meningitis, tuberculosis meningitis, central nervous system toxoplasmosis, HIV encephalopathy, or progressive multifocal leukoencephalopathy), or WHO “danger signs” of temperature >39 degrees Celsius, pulse >120 beats/minute, respiratory rate >30, or inability to walk unaided. Before undergoing any study procedure, all participants provided written informed consent.

### Randomization and Masking

Participants were randomized in a 1:1 ratio using a computer-generated random-number list in the GHESKIO Data Management Unit. A data manager who had no other involvement in study procedures conducted randomization by using a computer-number generator, using permuted blocks with variable block sizes from 2 to 6. The data manager transmitted participant allocation to the study physician. Participants, site personnel, and investigators were not masked to group assignment.

### Study Procedures

Participants were enrolled and randomized on the day of HIV diagnosis. In both groups, specimens were collected for creatinine, alanine transaminase (ALT), aspartate transaminase (AST), complete blood count (CBC) and CD4 count testing (FACS Count, Becton Dickinson, Franklin Lakes, NJ, USA); same-day results were not available. Participants also received a digital chest radiograph and provided a spot sputum specimen for Xpert Ultra (Cepheid, Sunnyvale, CA, USA), and liquid mycobacterial culture (Mycobacteria Growth Indicator Tube [MGIT], BACTEC, Beckton Dickinson, Franklin Lakes, NJ, USA). Specimens were refrigerated upon collection and transported to the central GHESKIO Biosafety Level 3 (BSL-3) laboratory across Port-au-Prince for testing. Key study interventions are illustrated in **Figure 1**.

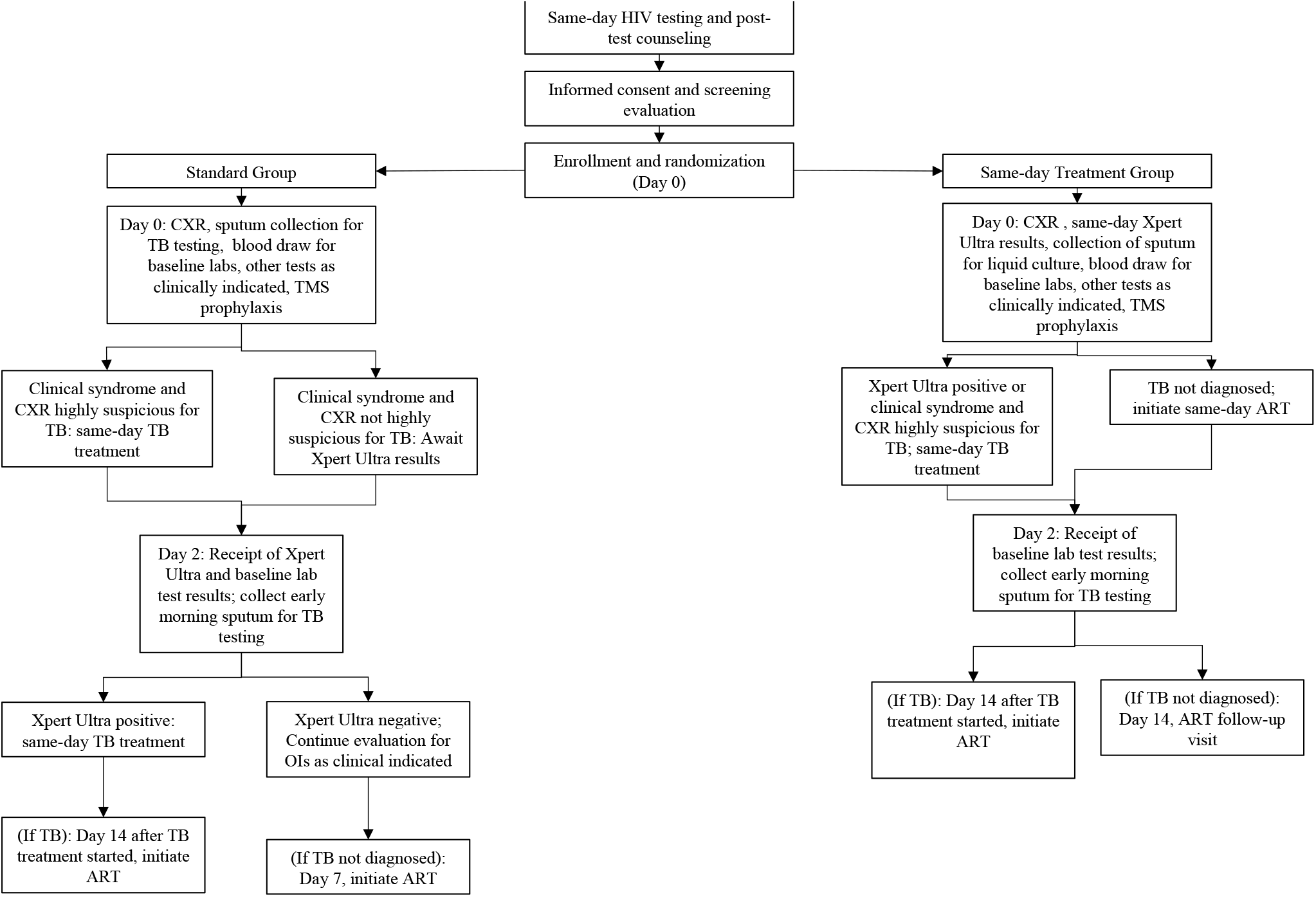

In the standard group, participants with a clinical syndrome and chest radiograph that were highly suspicious for TB initiated same-day TB treatment, without Xpert Ultra results. The remaining participants received neither ART nor TB medication on Day 0. All were discharged home with co-trimoxazole prophylaxis, as well as symptomatic therapy as clinically indicated. On Day 2, participants brought an early morning specimen to the laboratory for a second Xpert Ultra test and liquid mycobacterial culture and received the results of their first Xpert Ultra and baseline laboratory tests. Those with positive Xpert Ultra tests initiated same-day treatment; those with negative tests underwent continued evaluation for other opportunistic infections (OIs) as clinically indicated. On Day 7, those who had not initiated TB treatment started ART; initially this was Day 10, but in April 2018 it was changed to Day 7 to reflect changes in standard of care. Those with TB initiated ART after completing two weeks of TB treatment, in accordance with WHO guidelines (1).

The same-day group received same-day (Day 0) evaluation for TB, including Xpert Ultra results. This required immediate transport of sputum specimens to the central lab, prompt testing, and a phone call to study staff with test results. The study physician reviewed all clinical data and made a same-day determination about whether each participant had TB. All participants were discharged from the Day 0 visit with either TB medication or ART, as well as co-trimoxazole prophylaxis, and symptomatic therapy as clinically indicated. On Day 2, participants brought an early morning specimen to the laboratory for a second Xpert Ultra test and liquid mycobacterial culture, and received the results of their baseline laboratory tests, as for the standard group. Those with TB initiated ART after completing two weeks of TB treatment (1).

Other than the timing of availability of Xpert Ultra test results, and the timing of TB and ART treatment initiation, the same care was provided to both groups. First-line ART included efavirenz (EFV), tenofovir disoproxil fumarate (TDF), and lamivudine (3TC) until December 2018, when dolutegravir (DTG) became available. During 2019, all participants on EFV-based regimens were switched to DTG-based regimens, regardless of viral load results, in accordance with national guidelines. TB was treated with standard therapy in accordance with WHO guidelines (17). Prophylactic isoniazid was given to all participants who were not diagnosed with active TB. Participants with CD4 count <100 cells/mm^3^ were treated with 5 days of empiric azithromycin and those diagnosed with TB and CD4 count <100 cells/mm^3^ were prescribed prophylactic prednisone, in accordance with the REALITY and PredART study interventions (18, 19). Follow-up visits were scheduled at Weeks 4, 8, 12, 24, 36, and 48, as was standard of care during the study period. HIV-1 RNA testing was conducted at Weeks 24 and 48, in accordance with national guidelines. At every study visit, participants were queried about symptoms of IRIS using published criteria and definitions (20, 21).

Retention activities were similar to those provided for all PLWH at GHESKIO. Community health workers (CHWs) made reminder phone calls in advance of each visit, and after missed visits, with home visits for participants who could not be reached by phone. Participants received a transportation subsidy of 100 Haitian gourdes ($US 1.00) and a phone card (valued at 100 Haitian gourdes) at each visit. Participants were counseled about the importance of adhering to therapy, and of returning to clinic when they had symptoms.

### Outcomes

The primary outcome was the proportion of participants alive and in care with HIV-1 RNA <200 copies/mL at 48 weeks after enrollment, with intention to treat (ITT) analysis, with a pre-specified window of +/- 12 weeks; missing test results were considered virologic failures. Secondary outcomes included 48-week HIV-1 RNA cut-offs of <50 copies/mL and <1000 copies/mL, all-cause mortality, adherence as measured by medication possession ratio (proportion of days ART was dispensed within the study period), virologic failure with switch to second-line ART, sensitivity of spot and early morning Xpert Ultra, TB diagnosis after ART initiation, and incidence of IRIS. Bacteriologically confirmed (BAC+) TB required confirmation by molecular testing or culture. Empiric TB was defined as symptoms and chest radiograph consistent with TB, without BAC+ confirmation. Participants were considered retained if they attended the 48-week visit. LTFU was defined as lost to care without known death; those who returned for at least one visit after missing the 48-week visit were classified as returned to care. Deaths were defined by family report; death certificates are not available in Haiti. Transfer was defined as documented transfer to an outside clinic.

### Statistical Analysis

A target sample size of 388 was calculated to provide at least 80% power to detect an absolute difference of 14% in the proportion of participants with 48-week HIV-1 RNA <200 copies/mL (51% in the standard and 65% in the same-day treatment group), assuming a (two-sided) significance level of 0.05, using the Chi-square test. The target sample size was inflated to 600 to provide 80% power to detect an absolute difference in a secondary outcome of 48-week mortality of 6% (10% in the standard and 4% in the same-day treatment group), assuming a LTFU rate of 5% and significance level of 0.05.

Baseline characteristics were summarized using simple frequencies and proportions for categorical variables and medians with interquartile ranges (IQRs) for continuous variables by treatment group. We compared the proportion of participants with 48-week HIV-1 RNA <200 copies/mL (primary endpoint) and binary secondary outcomes using the Chi-square test (Fisher exact test was used instead for sparse data). For all comparisons, we presented unadjusted risk difference (RD) with 95% confidence interval (CI) and p-value.

For all analyses, an intention-to-treat approach was used, in which all participants were analyzed according to their assigned group. We did all analyses with SAS version 9.4. The study was registered with ClinicalTrials.gov NCT03154320. An independent data safety and monitoring board (DSMB) met twice per year to review study progress and participant safety.

## RESULTS

A total of 1738 adults (age ≥18 years) without known current TB treatment or pregnancy were diagnosed with HIV from November 6, 2017, to January 16, 2020. Of these, 669 (38.5%) reported at least one TB symptom during routine symptom screening and were informed about the study. Ninety-three patients were excluded prior to screening because they were not interested in study participation (n=50), not accepting of HIV diagnosis (n=23) or had a severe untreated co-morbidity (n=20). Of the 576 patients referred for screening, 500 were enrolled and randomized (n=250 per group) – see **Figure 2**, CONSORT Diagram. Enrollment was stopped on January 16, 2020, at the recommendation of the DSMB, because the target enrollment for the primary outcome (n=388) had already been surpassed, with numerically superior outcomes in the standard group. All 250 participants in each group were included in the analysis.

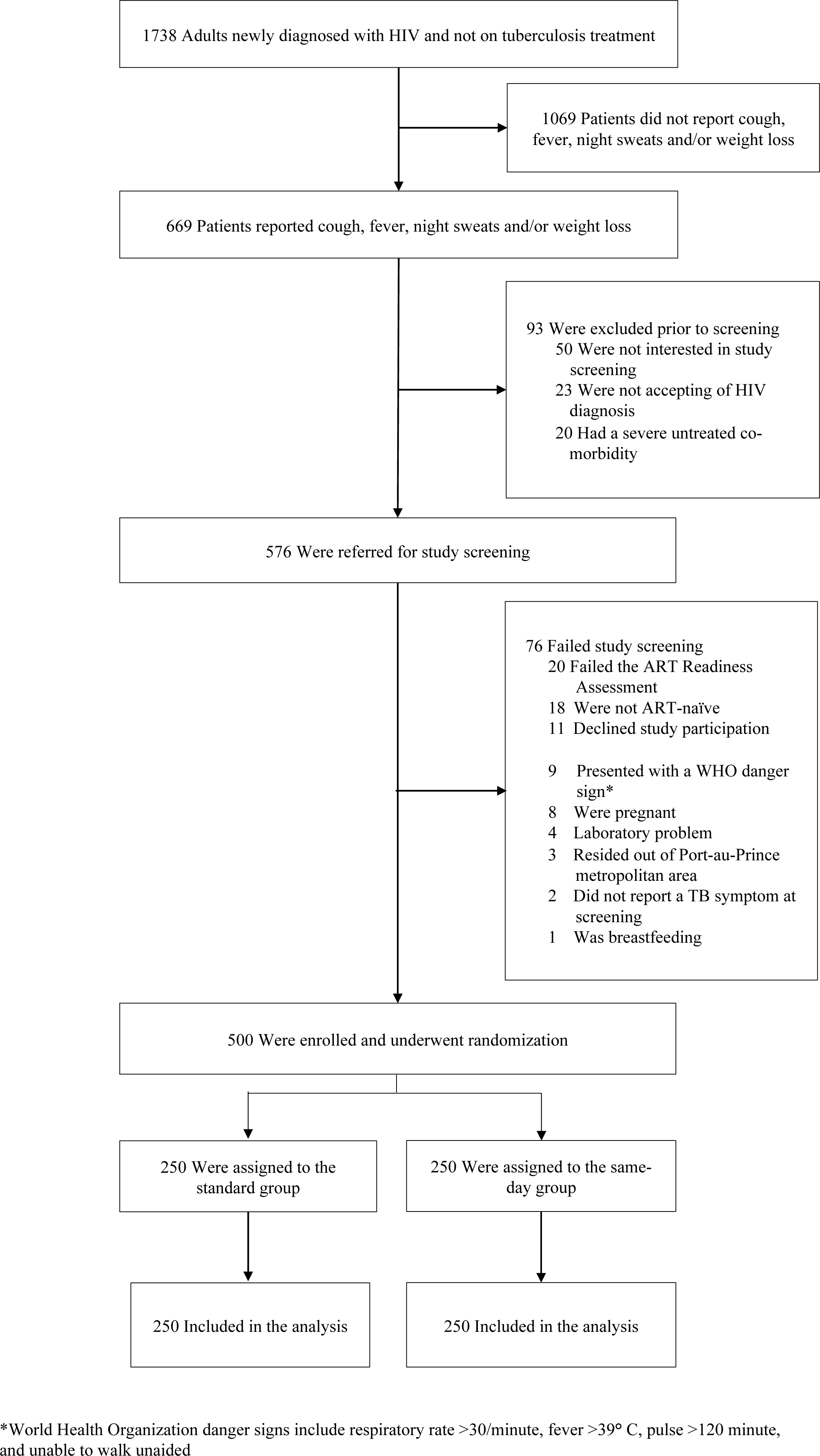

The median age of enrolled participants in the total cohort was 37 years (IQR: 30, 45), 234 (46.8%) were female, and 372 (74.4%) were living on <$US 1.00 per day (see **Table 1**). Two hundred (40.0%) participants reported cough, 194 (38.8%) reported fever, 72 (14.4%) reported night sweats, and 491 (98.2%) reported weight loss at study screening. Median body mass index (BMI) was 20.6 (IQR: 18.7, 22.9) and median CD4 count was 274 (IQR: 128, 426); 101 (20.2%) had CD4 count <100 cells/mm^3^. Forty (16.0%) participants in the standard group, and 48 (19.2%) in the same-day treatment group were diagnosed with baseline TB. Of these, 32 (80.0%) in the standard and 36 (75.0%) in the same-day treatment groups had BAC+ TB.

**Table 1.** Baseline Characteristics of Study Participants by Group.

|  | **Standard Group**<br>**(n=250)** | **Same-Day Treatment**<br>**Group (n=250)** |
| --- | --- | --- |
| Female – no. (%) | 117 (46.8) | 117 (46.8) |
| Age at enrollment (years) – median (IQR) | 37.0 (30.0, 45.0) | 36.0 (30.0, 45.0) |
| Income – no. (%) |  |  |
| <\$US 1.00 per day | 190 (76.0) | 182 (72.8) |
| ≥\$US 1.00 per day | 58 (23.2) | 65 (26.0) |
| Missing or unknown | 2 (0.8) | 3 (1.2) |
| Education level – no. (%) |  |  |
| None | 46 (18.4) | 38 (15.2) |
| Some primary | 95 (38.0) | 101 (40.4) |
| At least some secondary | 107 (42.8) | 107 (42.8) |
| Missing or unknown | 2 (0.8) | 4 (1.6) |
| Marital status – no. (%) |  |  |
| Single | 64 (25.6) | 70 (28.0) |
| Married | 137 (54.8) | 124 (49.6) |
| Formerly married | 47 (18.8) | 53 (21.2) |
| Missing or unknown | 2 (0.8) | 3 (1.2) |
| Tuberculosis symptoms – no. (%) |  |  |
| Reported cough | 100 (40.0) | 100 (40.0) |
| Reported fever | 96 (38.4) | 98 (39.2) |
| Reported night sweats | 34 (13.6) | 38 (15.2) |
| Reported weight loss | 243 (97.2) | 248 (99.2) |
| Body mass index – median (IQR) | 21.0 (18.8, 23.5) | 20.3 (18.5, 22.7) |
| CD4 count (cells/mm<sup>3</sup>) – median (IQR) | 293 (157, 447) | 228 (114, 398) |
| CD4 count category – no. (%) |  |  |
| <100 cells/mm<sup>3</sup> | 43 (17.2) | 58 (23.2) |
| 100 to 349 cells/mm<sup>3</sup> | 109 (43.6) | 116 (46.4) |
| 350 to 499 cells/mm<sup>3</sup> | 48 (19.2) | 36 (14.4) |
| ≥500 cells/mm<sup>3</sup> | 47 (18.8) | 38 (15.2) |
| Missing | 3 (0.6) | 2 (0.4) |
| Hemoglobin (g/dl) – median (IQR) | 12.0 (10.2, 13.6) | 11.7 (10.0, 13.4) |
| Tuberculosis status at HIV diagnosis – no. (%) |  |  |
| Bacteriologically confirmed tuberculosis | 32 (12.8) | 36 (14.4) |
| Empirically diagnosed tuberculosis | 8 (3.2) | 12 (4.8) |
| Tuberculosis not diagnosed | 210 (84.0) | 202 (80.8) |

Baseline TB prevalence varied by reported symptoms. Among participants in both groups who reported cough, fever, night sweats, or weight loss, 35.0% (70/200), 33.0% (64/194), 44.4% (32/72), and 17.3% (85/491), respectively, were diagnosed with TB. Among participants in both groups who reported only weight loss, 3.8% (9/239) were diagnosed with TB.

In the standard group, 33 of 40 (82.5%) participants diagnosed with baseline TB initiated treatment prior to ART initiation; of these, 30 (90.1%) initiated TB treatment within seven days of enrollment, and all 33 initiated ART at a median time of 14 days (IQR: 13, 14) after TB treatment. Seven (17.5%) participants (with negative Xpert Ultra and positive mycobacterial cultures) initiated ART prior to TB diagnosis; TB treatment was initiated within a range of 14 to 128 days after ART. Among the 210 participants who were not diagnosed with baseline TB, 205 (97.6%) initiated ART at a median time of seven days (IQR: 7, 10). Of these, 110 (53.7%) initiated ART within seven days, 83 (40.5%) from eight to 14 days and 12 (5.8%) >14 days after HIV diagnosis. Among all participants randomized to the standard group (regardless of TB status), ART was initiated a median of nine days (IQR: 7, 12) after enrollment.

In the same-day treatment group, 37 of 48 (77.1%) participants diagnosed with baseline TB initiated TB treatment on the day of HIV diagnosis; of these, 36 (97.3%) initiated ART at a median time of 14 days (IQR: 14, 15) after TB treatment, and one was LTFU prior to starting ART. Eleven (22.9%) participants initiated ART prior to TB treatment. Of these, five had negative spot but positive early morning Xpert Ultra tests and initiated TB treatment within a range of seven to 18 days after ART; six had negative Xpert Ultra tests, but positive mycobacterial cultures, and initiated TB treatment within a range of 37 to 89 days after ART. All 202 participants who were not diagnosed with baseline TB initiated ART on the day of HIV diagnosis. Among all participants randomized to the same-day treatment group (regardless of TB status), ART was initiated at a median time of 0 days (IQR: 0, 0) after enrollment.

Among the 250 participants in the standard group, 229 (91.6%) were alive and retained in care at 48 weeks after enrollment, and 220 (88.0%) received HIV-1 RNA testing with the 48-week visit window; of these, 168 (76.4% of those tested; 67.2% of those randomized) had HIV-1 RNA <200 copies/mL (see **Table 2**). Among the 250 participants in the same-day treatment group, 218 (87.2%) were alive and retained in care at 48 weeks after enrollment, and 211 (84.4%) received HIV-1 RNA testing within the 48-week visit window; of these, 152 (72.0% of those tested; 60.8% of those randomized) had HIV-1 RNA <200 copies/mL. There was no difference in the primary outcome (alive and retained in care with 48-week HIV-1 RNA <200 copies/mL) between the same-day and standard groups (risk difference -0.06; 95% CI: -0.15, 0.02; p=0.14).

**Table 2.** Study Outcomes by Group.

| Outcome | Standard Group<br>(n=250) | Same-Day Treatment<br>Group (n=250) | Risk Difference (95%<br>CI) | p-value |
| --- | --- | --- | --- | --- |
| **Primary Outcome** |  |  |  |  |
| 48-week HIV-1 RNA <200 copies/mL* | 168 (67.2) | 152 (60.8) | -0.06 (-0.15, 0.02) | 0.14 |
| **Secondary Outcomes** |  |  |  |  |
| 48-week HIV-1 RNA <50 copies/mL* | 159 (63.6) | 137 (54.8) | -0.09 (-0.17, -0.002) | 0.045 |
| 48-week HIV-1 RNA <1000 copies/mL* | 176 (70.4) | 159 (63.6) | -0.07 (-0.15, 0.01) | 0.11 |

| *Forty-Eight Week Outcomes\*\** |  |  |  |  |
| --- | --- | --- | --- | --- |
| Retained in care* | 229 (91.6) | 218 (87.2) | -0.04 (-0.10, 0.01) | 0.11 |
| Died | 6 (2.4) | 9 (3.6) | 0.01 (-0.02, 0.04) | 0.43 |
| Lost to follow-up | 10 (4.0) | 14 (5.6) | 0.02 (-0.02, 0.05) | 0.40 |
| Missed 48-week visit due to gap in care,<br>with subsequent return to care | 4 (1.6) | 6 (2.4) | 0.01 (-0.02, 0.03) | 0.52 |
| Transferred | 1 (0.4) | 3 (1.2) | 0.01 (-0.01, 0.02) | 0.62 |

\*Nine participants in the standard treatment and seven in the same-day group attended 48-week visit without HIV-1 RNA testing; \*\* These are mutually exclusive outcomes which include all 500 randomized participants.

Among the 220 participants in the standard and 211 in the same-day treatment group who received 48-week viral load testing, 159 in the standard group had HIV-1 RNA <50 copies/mL (72.3% of those tested; 63.6% of those randomized), compared with 137 in the same-day group (64.9% of those tested; 54.8% of those randomized). The same-day group was less likely than the standard group to achieve 48-week HIV-1 RNA <50 copies/mL (54.8% vs. 63.6%; RD: -0.09; 95% CI: -0.17, -0.002) – see **Table 2**. With an HIV-1 RNA cut-off of 1000 copies/mL, 176 in the standard group (80.0% of those tested; 70.4% of those randomized) and 159 in same-day treatment group (75.4% of those tested; 63.6% of those randomized) achieved viral suppression; there was no difference between the same-day and standard groups (63.6% vs. 70.4%; RD: -0.07; 95% CI: -0.15, 0.01).

In the standard group, 21 participants (8.4%) were not retained in ART care at 48 weeks after enrollment. A total of six (2.4%) died, 10 (4.0%) were LTFU, four (1.6%) had a gap in care during the 48-week visit window with subsequent return to care, and one (0.4%) was transferred (see **Table 2**). In the same-day treatment group, 32 (12.8%) participants were not retained in ART care at 48 weeks after enrollment. A total of nine (3.6%) died, 14 (5.6%) were LTFU, six (2.4%) had a gap in care during the 48-week visit window with subsequent return to care, and three (1.2%) were transferred. There were no differences between groups in retention (87.2% vs. 91.6%; RD: -0.04; 95% CI: -0.10, 0.01) or mortality (3.6% vs. 2.4%; RD: +0.01; 95% CI: -0.02, 0.04).

Three (1.2%) participants in the standard group were diagnosed with incident TB; two of these were treated empirically for pulmonary TB, and one for extrapulmonary TB. One (0.04%) participant in the same-day group was diagnosed with incident TB (Xpert Ultra/culture-positive tuberculous lymphadenitis). No participant in either group was diagnosed with either unmasking or paradoxical IRIS during the first 12 weeks after ART initiation.

Among the six participants in the standard group who died during the study period, two deaths were attributed to incident TB, one to mycobacterial avium complex, one to advanced AIDS with OI of unclear etiology, one to tear gas inhalation during a street protest, and one cause of death was unknown. Among the nine participants who died in the same-day treatment group; three were attributed to advanced AIDS with OI of unclear etiology, one to advanced AIDS and gastroenteritis, one to Kaposi’s sarcoma diagnosed after enrollment, one to cancer, one to renal failure, and two were unknown.

Among the participants who received 48-week viral load testing, 61.8% (136/220) in the standard and 65.9% (139/211) in the same-day group had adherence ≥90% by pharmacy refill; there were no differences in adherence between the two groups (RD: 0.04; 95% CI: -0.05, 0.13). Among participants with 48-week HIV-1 RNA ≥200 copies/mL, adherence was <90% among 76.9% (40/52) in the standard and 57.6% (34/59) in the same-day treatment group.

Among the 245 participants in the standard group who initiated ART, 70 (28.6%) received only an EFV-based regimen, 83 (33.9%) initiated EFV and switched to a DTG-based regimen due to drug availability, 90 (36.7%) received only a DTG-based regimen, and two (0.8%) initiated EFV and switched to a second-line PI-based regimen. Among the 249 participants in the same-day treatment group who initiated ART, these numbers were 73 (29.3%), 75 (30.1%), 94 (37.8%), and seven (2.8%) respectively. There was no difference between groups in switches to second-line ART (2.8% vs. 0.8%; RD: 0.02; 95% CI: -0.004, 0.04; p=0.18).

## DISCUSSION

The results of this randomized trial show that in patients with TB symptoms at initial HIV diagnosis, same-day treatment is not associated with superior outcomes. The findings of this study do not confirm our hypothesis that same-day TB assessment and same-day treatment with TB medication or ART would improve retention with viral suppression in this patient population. All primary and secondary pre-specified outcomes indicate that same-day treatment is not superior to standard care. However, an important caveat is that treatment was provided rapidly in the standard group, with median time to ART initiation of only 7 days in those not diagnosed with TB, which meets WHO criteria for rapid ART initiation (6). Our findings indicate that either strategy is appropriate, but the standard group, which included a short delay in ART initiation for TB assessment, is logistically simpler.

In prior randomized trials which found superior rates of retention and viral suppression with same-day ART, including a study conducted at our site in Haiti, the majority of standard group participants delayed ART initiation for weeks to months after HIV diagnosis (11, 12, 14, 22). In contrast, in this study delays in treatment initiation were minimized for all participants, reflecting current practice, and the standard group achieved rates of viral suppression that were numerically similar or higher than those of the same-day ART groups in previous trials, which ranged from 44% to 64% (11-14).

In contrast to our approach, in which all participants received at least one Xpert Ultra result prior to ART initiation, participants in the intervention group in the Simplified Algorithm for Treatment Eligibility (SLATE) II study with nonserious TB symptoms and a negative urine point-of-care lipoarabinomannan antigen of mycobacteria (LAM) test were eligible for same-day ART, with next-day tracing to contact those with positive Xpert MTB/RIF results (13). Rates of rapid ART initiation and 8-month retention were higher in the intervention group, with no reported serious TB-related adverse events; however, only three of the seven confirmed TB cases in the intervention group initiated same-day ART.

Eighteen percent of participants in our study cohort were diagnosed with TB at baseline. Though a small number started ART prior to TB diagnosis, and about 20% presented with CD4 counts <100 cells/mm^3^, no cases of IRIS were detected during the first 12 weeks after ART initiation (20, 21). This provides some reassurance about initiating ART after only a single negative Xpert Ultra test. However, participants with symptoms and chest radiographs that were highly suggestive of TB were initiated on TB treatment, regardless of sputum testing results. Furthermore, prophylactic prednisone was prescribed for participants with TB and CD4 count <100 cells/mm.^3^

The implications of our study findings are that the pendulum has potentially swung too far in favor of same-day ART. Within the model of differentiated service delivery, patients do not all have the same needs. Same-day ART is ideal for many patients, but may not be necessary for those with TB symptoms, in whom a short delay in treatment initiation does not appear to compromise outcomes. The provision of same-day Xpert MTB/RIF results is logistically complicated, particularly in large sites such as ours, with centralized laboratories. An alternative approach would be initiating ART while Xpert results are pending, but we agree with the systematic review which concluded that there is insufficient evidence to recommend this strategy in settings with high burdens of TB (9). Only a small number of patients with TB have been included in same-day ART studies, and nearly all received EFV-based regimens (9, 12-14, 23). However, DTG is associated with faster improvements in immune function. ART-mediated immune restoration may cause pulmonary dysfunction, even in patients on TB treatment without clinical evidence of TB-IRIS; initiating DTG-based ART in the presence of untreated TB could potentially provoke even greater declines in pulmonary function (7, 8). Post-TB lung disease may contribute to the higher long-term mortality reported in patients cured of TB, compared with control populations (24, 25).

It is critical to emphasize that the GHESKIO approach includes more than just ART initiation, as other successful programs have also described (11-14, 22, 23, 26). GHESKIO staff are trained in building a relationship with patients, including creating a welcoming environment, minimizing stigma, and explaining the purpose of testing for TB and other conditions, and the importance of treatment adherence. Moreover, all patients who are diagnosed with HIV at GHESKIO – regardless of study participation – are prescribed co-trimoxazole prophylaxis at a minimum on the day of HIV diagnosis. GHESKIO also provides medications free of charge for relief of common symptoms, such as cough and fever. In this way, every patient who is newly diagnosed with HIV leaves GHESKIO with at least one medication and support for their treatment adherence – as well as a hopeful message that HIV is a treatable disease.

Nearly two-thirds of the total study cohort achieved the primary outcome of retention in care with 48-week viral suppression. Retention rates were high, but viral suppression rates were sub-optimal in both groups, though similar to global estimates from the International epidemiology Databases to Evaluate AIDS (IeDEA) consortium (27). It is noteworthy that Haiti suffered from severe political instability, civil unrest, and gang-related violence throughout the study period and there were added disruptions from the COVID-19 pandemic (28). In this context, it was not always feasible for participants to receive ART refills in a timely manner. Furthermore, first-line ART included an EFV-based regimen for the first year of study enrollment, and EFV resistance has been reported in over 20% of ART-naïve patients at GHESKIO (29).

Our study was conducted in a large urban clinic, which may limit the generalizability of our findings. It is also noteworthy that we utilized a more aggressive diagnostic approach for TB than is generally provided in usual care, which may have contributed to our lack of TB-IRIS. Moreover, all participants received medication for prophylaxis of OIs or symptomatic relief on the day of HIV diagnosis, which may have contributed to our high retention rate in the standard group. However, we note the prescription of non-ART medications on the day of HIV diagnosis is common practice in many settings.

In conclusion, our results indicate that same-day treatment is not associated with superior outcomes, compared with standard care, for patients who present with TB symptoms at HIV diagnosis. Our findings suggest that a short delay to assess for TB does not appear to compromise treatment outcomes. This precludes the need to initiate ART prior to completion of initial TB testing, which may result in a substantial proportion of co-infected patients initiating ART before TB treatment.

## Data Availability

All data produced in the present study are available upon reasonable request to the authors

## Acknowledgements

We thank the patients who participated in this study, and all of the GHESKIO staff who cared for them. We also thank the members of the data safety monitoring board, the community advisory board, and the ethics committees for their expertise and advice.

